# Challenges associated with diagnostic exome sequencing in liver diseases

**DOI:** 10.1101/2022.06.21.22276400

**Authors:** Xiao-Fei Kong, Kelsie Bogyo, Sheena Kapoor, Emily E. Groopman, Amanda Thomas-Wilson, Enrico Cocchi, Hila Milo Rasouly, Beishi Zheng, Siming Sun, Junying Zhang, Mercedes Martinez, Jennifer M Vittorio, Lorna M. Dove, Maddalena Marasa, Timothy C. Wang, Elizabeth C. Verna, Howard J. Worman, Ali G. Gharavi, David B. Goldstein, Julia Wattacheril

## Abstract

Exome sequencing (ES) has been used in a variety of clinical settings but there are limited data on its utility for diagnosis and/or prediction of monogenic liver diseases. We analyzed ES data in 758 patients with liver diseases., We developed a curated list of 502 genes for monogenic disorders associated with liver phenotypes and analyzed ES data for these genes in 758 patients with chronic liver diseases (CLD). For comparison, we examined ES data in 7,856 self-declared healthy controls (HC), and 2,187 patients with chronic kidney disease (CKD). Candidate pathogenic (P) or likely pathogenic (LP) variants were initially identified in 19.9% of participants, most of which were attributable to previously reported pathogenic variants with implausibly high allele frequencies. After variant annotation and stringent filtering based on population minor allele frequency, we detected a significant enrichment of P/LP variants in the CLD cohort compared to the HC cohort (*X*^2^ test OR: 5.00, 95% CI:3.06-8.18,, p-value=4.5 e-12). A second-level manual annotation was necessary to capture true pathogenic variants that were removed by stringent allele frequency filters. After these sequential steps, the diagnostic rate of monogenic disorders was 5.7% in the CLD cohort, attributable to P/LP variants in 25 genes. We also identified concordant liver disease phenotypes for 15/22 kidney disease patients with P/LP variants in liver genes, mostly associated with cystic liver disease phenotypes. Clinically confirmed sequencing results had many implications for clinical management, including familial testing for early diagnosis and management, preventative screening for associated comorbidities, and in some cases for therapy. Exome sequencing provided a 5.7% diagnostic rate in CLD patients and required multiple rounds of review to reduce both false positive and false negative findings. The identification of concordant phenotypes in many patients with P/LP variants and no known liver disease also indicates a potential for predictive testing for selected monogenic liver disorders.

## Introduction

Liver disease accounts for approximately 2 million deaths per year worldwide. In the United States, the mortality rate for chronic liver diseases (CLD) increased 31% from 2000 to 2015, making it the fifth leading cause of death in 2017 for persons aged 45-64 years ^1^. The history of liver genetic diseases dates back to 1865-1890 when Triouseau and von Recklinghausen described hemochromatosis ^2^. The cloning, mapping, and functional characterization of homeostatic iron regulator (*HFE*) gene in the 1990s paved the way for molecular diagnosis of hemochromatosis ^3^. The advent of next-generation sequencing (NGS) approaches have led to the discovery of genetic disorders causing liver disease phenotypes such as fibrolamellar hepatocellular carcinoma ^4^, recurrent acute liver failure^5–7^, or idiopathic non-cirrhotic portal hypertension^8,9^. These findings demonstrate the power of NGS for identifying novel genetic forms of liver diseases.

NGS has been successfully deployed in clinical care to diagnose monogenic forms of neurologic, developmental, cardiac or renal disorders ^10,11^. While genetic testing of single genes or small gene panels has been used for some suspected hereditary liver diseases ^14–18^, NGS approaches have not been widely adopted into the routine evaluation of liver disease. As sequencing costs decline and clinical utility is demonstrated, a standardized genetic diagnostic pipeline for liver disease could benefit patients and clinicians, enabling efficient clinical diagnoses and early recognition of rare genetic disorders that may manifest as a common liver phenotype and may not be recognized based on their clinical workup^12^. In this paper, we outline an analytic approach and conduct a clinical sequence interpretation for ES data from 10,801 individuals, including 758 patients with CLD as encountered at various stages of their diagnostic workflow. Here we present the diagnostic utility of ES for liver diseases, highlight special considerations and elaborate on the potential for misclassification in the genetic workup for liver diseases.

## Results

### 1. Characterization of 502 liver genes with Mendelian hepatobiliary disorders

In a comprehensive search for Mendelian genetic disorders with any liver abnormalities prompting clinical referral to a hepatologist, we manually curated a total of 959 genes. Of these, 502 had a confirmed abnormal liver disease phenotype, with 193 genetic disorders having primarily liver disease. For example, *ABCB11* or *ATP8B1* causing progressive familial intrahepatic cholestasis; other genes might lead to liver abnormalities that present clinically as a secondary cause (**Supplementary Figure 1** and **Supplementary Appendix 1**). We then annotated inheritance modes and detailed clinical phenotypes related to these 502 genes. In total, 75% of genes were associated with a recessive mode of inheritance (363 AR and 15 XLR). Sixty-two autosomal genes could result in both dominant and recessive disorders and 62 other genes associated with exclusively dominant disorders (61 AD and 1 XLD) (**Supplementary Table 3**). The most common clinical presentation of Mendelian hepatobiliary disorders was hepatomegaly, manifesting in 236/502 disorders (47%) Other common clinical manifestations included metabolic disease (25%), liver fibrosis or cirrhosis (25%), elevated hepatic transaminase level (20%), and cholestasis (19%) (**Figure 1A**). Most of the genes (298/502, 59%) were associated with a developmental or congenital disorder with liver manifestations (**Supplementary Table 3**). The 62 genes exclusively associated with dominant inheritance showed significantly higher pLI (**Figure 1B**) and missense Z scores (**Figure 1C**) compared to the 378 genes associated with recessive diseases or the 62 genes associated with both dominant and recessive inheritance. Based on these results, genes associated with both dominant and recessive disorders were analyzed under the recessive inheritance model.

**Figure 1:**
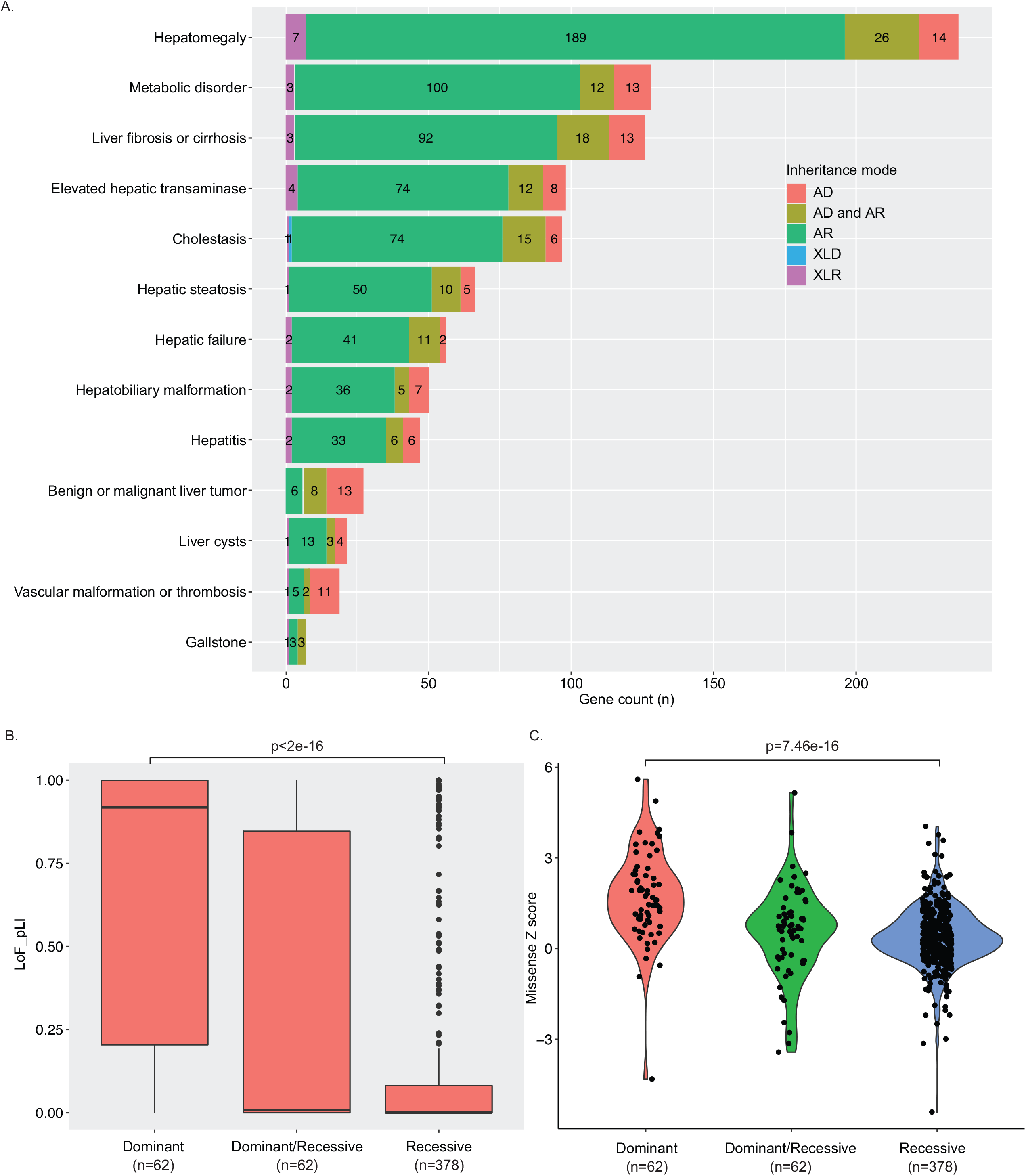
A summary of liver phenotypes in Mendelian genetic disorders. A) Inheritance mode, annotated clinical liver phenotypes, and biological effects of 502 genes related to Mendelian disorders. The liver phenotypes and inheritance were curated based on OMIM, ClinGen, and a literature search. AD: autosomal dominant disorder; AR: autosomal recessive disorder; XLD: X-linked disorder; XLR: X-linked recessive disorder; AD and AR: Genes with both autosomal dominant and autosomal recessive inheritance were reported. The right lower box showed the numbers of genes with corresponding biological effects and inheritance mode. B) Box plot of pLI scores of 502 genes in three groups based on inheritance mode. The dark line inside the box represents the median of pLI score. The top of box is 75% and bottom of box is 25%. The endpoints of the lines are at a distance of 1.5*IQR, where inter quartile range is the distance between 25th and 75th percentiles. The points outside the whiskers are marked as dots and are considered as extreme points. C) Violin plot of missense Z scores of 502 genes in three different groups based on inheritance mode. P-values in B and C for differences between dominant and recessive genes were determined using ANOVA.

### 2. Assessment of the frequency of candidate pathogenic/likely pathogenic variants

To investigate the prevalence of candidate pathogenic variants in the liver genes, we analyzed ES data from 10,801 individuals, agnostic to the clinical phenotype. Based on automated filtering, we initially identified an equal distribution of candidate pathogenic variants across the three cohorts: 1,567 (20.2%) in healthy controls, 416 (19.0%) in the CKD cohort, and 159 (21%) in the CLD cohort (**Supplement Figure 2A and 2B, Supplementary Table 4**). This implausibly high frequency of variants for monogenic liver disorders suggested variant misclassification. Consistent with this conjecture, an analysis of CADD score and the maximal MAF from the ExAC and gnomAD indicated that many of these variants had implausibly high allele frequencies to be disease causing and had been erroneously reported as pathogenic prior to the availability public variant databases (**Supplement Figure 2C**). We next used the maximal MAF to filter these variants, followed by manual review of 403 variants. (**Supplementary Table 5, Supplementary Figure 2A and 2B**)^30,34,35^. This resulted in 112 variants being classified as either P/LP based on ACMG-AMP classification (including 78 PTVs, **Supplement Figure 2D**), detected across 45 genes in a total of 100 individuals (0.93% of three cohorts). Subsequent to this filtering and manual annotation process, the prevalence of these P/LP variants significantly differed between healthy controls (51/7856, 0.65%), patients with CKD (25/2187, 1.14%), and patients with CLD (24/758, 3.17%) (*X*^*2*^ test OR: 5.00, 95%CI:3.06-8.18, p-value=4.55e - 12, **Figure 2A** and **Supplementary 2E)**.

**Figure 2:**
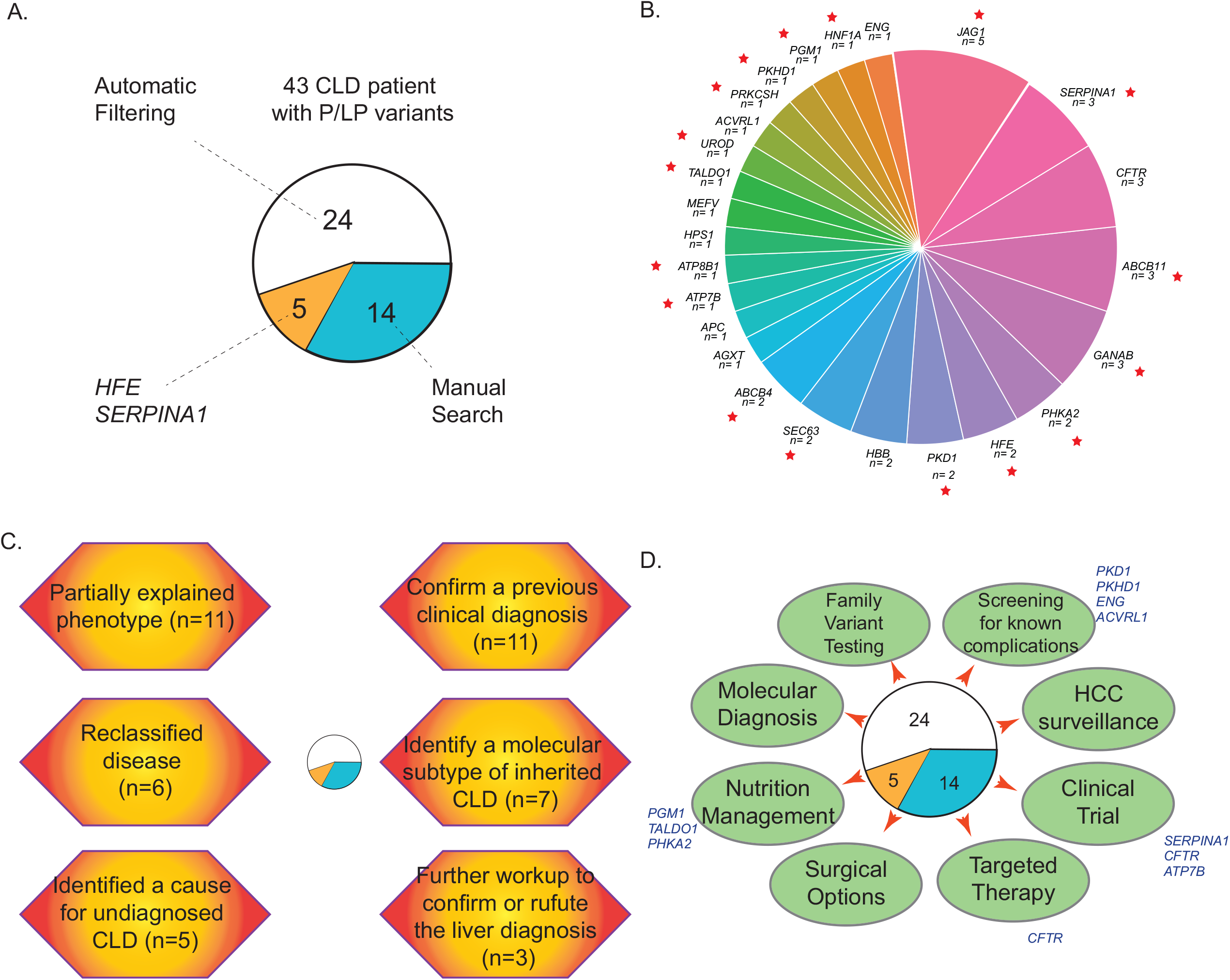
Genetic diagnoses and clinical implications of ES findings in the liver disease cohort. A) A total of 43 CLD patients with P/LP variants from three searching approaches; B. A total of 25 genetic disorders were found in the CLD cohort. Red star indicated the genetic disorders causing primarily liver diseases. A genetic characterization (C.) and clinical implication (D.) with the findings.

**Figure 3:**
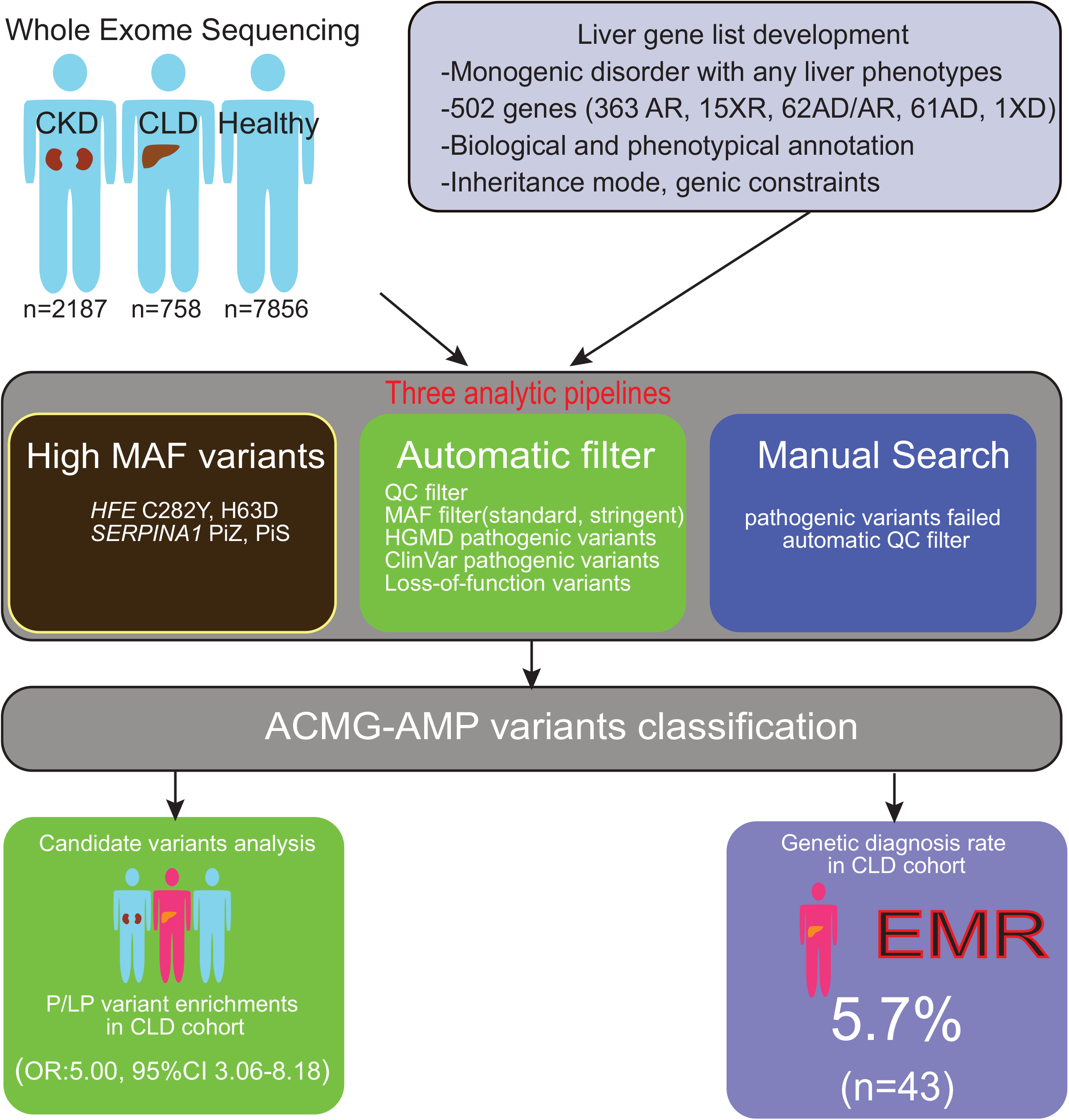
A summary of the genetic analytic strategy and outcomes for liver diseases.

### 3. Second-level annotation of the CLD cohort identifies additional pathogenic variants

To maximize the identification of diagnostic variants in the CLD cohort, we performed a second-level manual assessment, using the more relaxed sequence quality thresholds which we had previously deployed to optimize diagnostic yield in other cohorts ^31,38^. This second-level analysis led to the identification of 16 additional diagnostic variants that explained the liver phenotypes in 14 additional patients (13 genes, **Figure 2A**). All 16 variants were missed because of the high stringency sequence quality thresholds and were all confirmed by Sanger sequencing. In addition, we evaluated five well-known pathogenic variants or risk alleles for liver disease that have a MAF above 1%: *HFE* C282Y and H63D, *SERPINA1*E264V (Pi*S) and E342K (Pi*Z). We found two patients with P/LP variants in *HFE* (one with a homozygous *HFE* C282Yvariant, and one with an H63D/c.340+1G>A genotype, **Table 2**). Both had high serum iron transferrin saturation and ferritin levels, and clinical presentations consistent with hereditary hemochromatosis. For *SERPINA1*, three patients in the CLD cohort had a homozygous Pi*Z genotype, and all of them had a clinical diagnosis of alpha-1 anti-trypsin deficiency (**Table 2**). Altogether, this second level analysis increased the diagnostic yield in the liver cohort to 43/758 cases (5.7%, **Figure 2A**).

**Table 2:** *HFE* and *SERPINA1* variants in three cohorts

| Genotypes | HC<br>(n=7856) | CKD<br>(n=2187) | CLD<br>(n=758) |
| --- | --- | --- | --- |
| ***HFE* variants** |  |  |  |
| Homozygous C282Y | 2 | 1<sup>\$</sup> | 1<sup>\$</sup> |
| Homozygous H63D | 9 | 3<sup>&</sup> | 0 |
| Compound C282Y/H63D* | 60 | 11 | 2 |
| C282Y or H63D/PTV | 22 | 5 | 1<sup>\$</sup> |
| Total(n=117) | 93 (1.20%) | 20 (0.91%) | 4(0.53%) |
| ***SERPINA1* variants:** |  |  |  |
| Pi ZZ | 0 | 0 | 3<sup>\$</sup> |
| Pi MZ (n=222) | 138 (1.76%) | 49 (2.24%) | 15 (1.98%) |
| Pi SS | 1 | 0 | 0 |
| Pi MS(n=606) | 482 (6.14%) | 92 (4.21%) | 32 (4.22%) |
| Pi Z/ Pi S* (n=12) | 7 | 5 | 0 |

### 4. Genetic diagnoses and their clinical implications

Overall, we identified a total of 25 genetic disorders in the liver disease cohort, with Alagille syndrome, alpha-1 anti-trypsin deficiency, cystic fibrosis, and progressive familial intrahepatic cholestasis-2 detected in at least three patients each **(Figure 2B)**. There were no differences observed in sex, race, or ethnicity between the patients with or without a genetic diagnosis in the liver disease cohort. From a univariate analysis, younger age and the clinical diagnosis of congenital liver disorders, abnormally elevated serum transaminase activities due to unknown causes were associated with a higher rate of a genetic diagnosis (**Table 1**). We next performed a case-level review to assess concordance between genotype and phenotype. Among 43 liver disease patients with P/LP variants, we confirmed a previous clinical diagnosis for eleven, identified a genetic disease that partially explained the phenotype for eleven, reclassified disease for seven, identified a molecular subtype of inherited liver diseases for six, and identified a cause for undiagnosed liver diseases for five. We also recommended further workup in three patients to confirm or refute the liver diagnosis (**Figure 2C**). In addition, we examined the phenotypic concordance for the 25 kidney patients carrying P/LP variants in liver genes: 15/25 patients had a corresponding liver phenotype, which were mostly attributable to P/LP variants in genes like *PKD1*, MODY or ciliopathy genes causing both kidney and liver disease (**Supplement Appendix 2)**. Benefits of a genetic diagnosis included the ability to guide familial testing and obtain an early diagnosis of affected family members for 24 families, or to perform surveillance for known complications, such as brain aneurysms in individuals carrying a pathogenic variant in *PKD1*. Four patients with *HFE* and *PFIC2* will be followed clinically for progression to appropriate stages of disease for cancer screening. Patients with *PGM1* and *PHKA2* pathogenic variants, diagnostic of congenital disorders of glycosylation, can benefit from selective nutritional management. Other implications for better treatment include targeted therapy, clinical trials, or surgical options. For example, a review of clinicaltrials.gov identified 255 clinical trials are enrolling patients with monogenic forms of liver diseases identified in this study (**Figure 2D**).

**Table 1:**
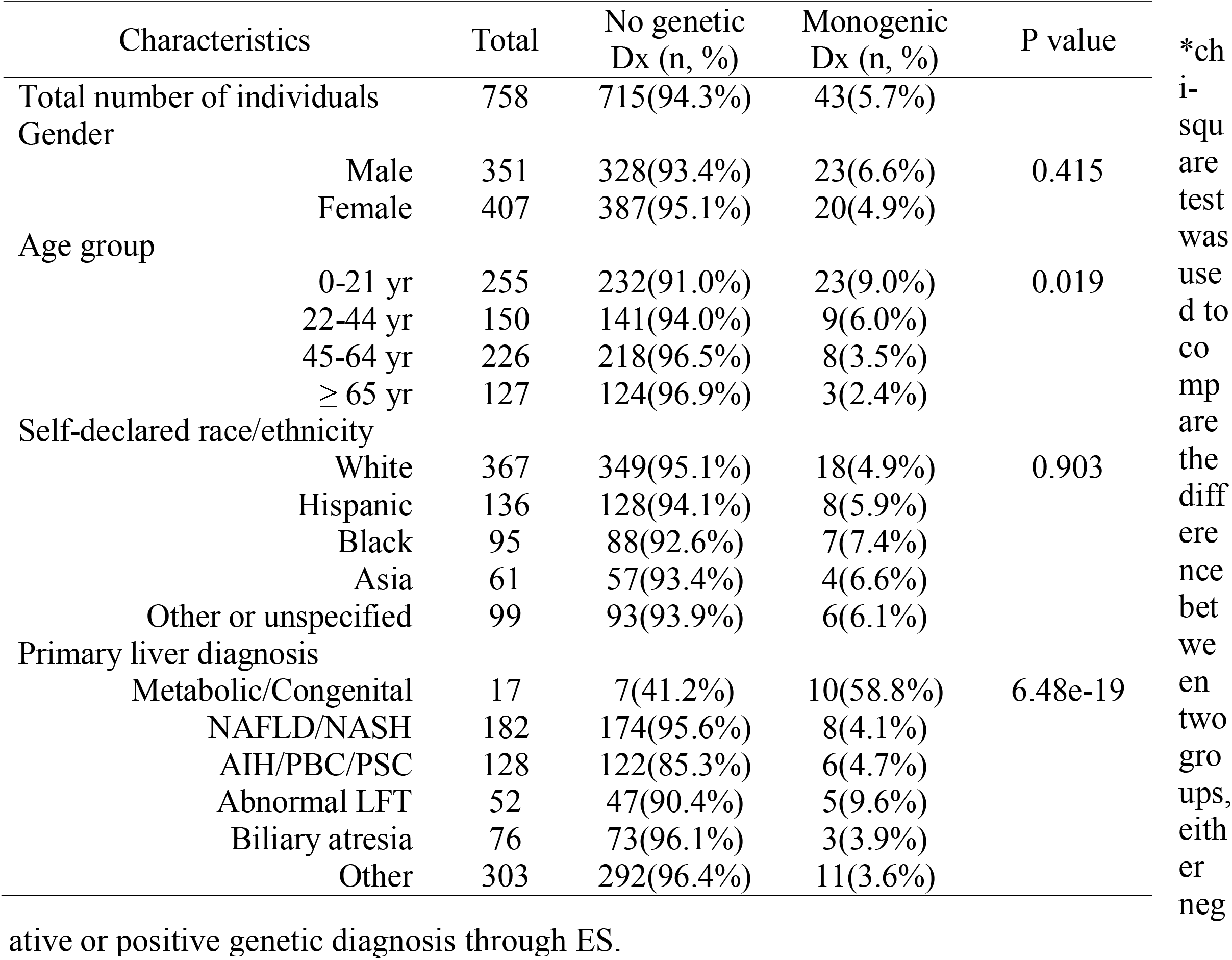
Clinical characteristics for monogenic diagnosis in the liver cohort from ES analysis

## Discussion

Our primary goal was to evaluate the utility of ES for diagnosis of liver disease. Currently available clinical genetic testing for heritable liver diseases exists and is mostly utilized in the pediatric populations. For instance, one lab provides a panel of 72 genes for well-defined monogenic liver diseases, especially cholestasis and biliary atresia^39^. To guide the ES analysis, we developed a list of 502 genes associated with a Mendelian disease with potential liver phenotypes. This work constitutes an initial attempt at a gene list for monogenic liver disease, but the list will have to be continuously annotated and updated to include new information about genes and variants. For example, we updated the list to include several genes (*TULP3*^40^, *KIF12, USP54*^41^, *KCNN3*^42,43^, *GIMAP5*^9^) which have been implicated in monogenic disorders associated with liver phenotypes during the performance of this study.

We also removed some genes which, in retrospect, did not have a secure causal relationship with CLD. In the future, the creation of a liver disease workgroup, for instance, under the ClinGen platform, will accelerate the development of a reference gene list for CLD.

The current challenge of genetic analysis is to determine the pathogenicity of variants. In this work, we focused on genes associated with monogenic disorders and omitted analysis of risk factors, such as *PNPLA3*. Consistent with prior studies of other genetic disorders, our variant level analyses indicated that many previously reported P/LP variants for liver diseases are too common to be pathogenic and are erroneously annotated in reference databases. We report liver disease genes with the most frequently encountered false-positive P/LP variants to help with the reannotation of reference databases (Supplementary Appendix 2). We also performed a manual annotation of the data, which confirmed that the application of hard filters for allele frequency and sequence quality may lead to the omission of true pathogenic variants. For example, in addition to *HFE* and *SERPINA1* pathogenic variants, two patients with progressive familial intrahepatic cholestasis type 3 carried an *ABCB4* Ala934Thr missense variant which has a MAF of 1.2% in African-American populations, but should be interpreted as a pathogenic variant. Thus, a balanced disease-specific approach was necessary for maximizing the diagnostic rate. A case-level review indicated that the genetic results were consistent with the clinical findings in the majority of liver and kidney disease cases, validating our approach. The genetic findings had many implications for diagnosis, risk stratification, surveillance, sometime for therapy, including potential eligibility for clinical trials. For the patients who did not have a concordant liver disease phenotype, the P/LP variants may be non-penetrant, disease may develop in the future, or the variant may be downgraded in the future based on evidence of non-pathogenicity. We note that our study is limited by the lack of clinical information for most self-reported healthy controls, which hampers our ability to determine the causality of P/LP variants in this cohort.

Altogether, our single-center study indicates a significant diagnostic utility for ES in the evaluation of patients with CLD. We note the pitfalls for diagnostic analysis based on hard filters, necessitating a more domain specific approach to variant annotation. Future studies will have to evaluate the diagnostic utility across multiple healthcare settings and prospectively demonstrate the impact of genetic testing on clinical decision-making, cost-effectiveness and genetically stratified clinical studies.

## Material and Methods

### Developing a list of monogenic disorders associated with liver phenotypes

We first worked on a gene list, or “liver gene list”, to identify genes causing monogenic diseases with a wide range of liver manifestations. We used Online Mendelian Inheritance in Man database (OMIM), Orphanet, and the Human Phenotype Ontology (HPO) database ^19^ to search for potential genes with Mendelian inheritance that have been associated with or shown to be causative in liver disease before December 2018. For the search, we used a total of 30 keywords or phrases (**Supplementary Figure 1**), then manually reviewed OMIM and related literature ^20–27^. We excluded: 1) genes not reported to be linked to any abnormal liver phenotypes; 2) genes within a locus reported from linkage analysis without any known pathogenic variants; 3) genes only discovered in GWAS but lacking any evidence for Mendelian inheritance; 4) genes with only somatic variants reported in abnormal liver phenotypes; 5) genes within a locus associated with abnormal liver phenotypes due to chromosomal abnormalities. The selected genes were annotated for biological functions, clinical liver presentations, and gene constraint score ^28^. We annotated the inheritance mode of liver genes based on OMIM and ClinGen then manually curated the list of genes by reviewing relevant literature. The current gene list is an initial attempt to catalog monogenic liver diseases and remains a work in progress. We anticipate that this list will require regular updates and curations and may serve as the basis for a reference liver gene list that can be curated by an expert group, such as the ClinGen^29^.

### Cohorts

We analyzed ES data obtained by sequencing of genomic DNA from 758 patients with CLD. We enrolled patients from both pediatric and adult liver clinics at Columbia University Irving Medical Center (CUIMC) who were interested in and able to consent to participating in genetics research, without setting inclusion or exclusion criteria (**Table 1**). In the CLD cohort, 53.7% of participants were female, and 33.6% of participants were under 22 years of age. 182 patients with CLD (24%) were diagnosed with nonalcoholic fatty liver disease (NAFLD) or nonalcoholic steatohepatitis (NASH), 128 patients (16%) with AIH or PBC or PSC, other patients with viral hepatitis (n=125), and alcoholic hepatitis (n=27) were also included. A few cases with acute liver failure (n=5), or hepatocellular carcinoma (HCC, n=3), or hepatoblastoma (n=7), or cardiogenic liver cirrhosis (n=9) were sequenced and analyzed altogether. A selection bias might occur as we attempt to enroll those who might have a genetic cause of liver diseases. A detailed clinical description is provided in **Supplementary Table 1** and **Supplementary Table 2**. In additional, two control cohorts were used to evaluate the gene-list based ES analysis, including 7,856 self-identified healthy individuals and 2,187 patients from CUIMC with chronic kidney disease (CKD) (**Supplementary Table 1**)^30,31^. The CKD cohort was included because we had access to health records through CUIMC, enabling us to evaluate the penetrance of monogenic liver disorders in a cohort not ascertained for liver disease. Informed consent in writing was obtained from each patient and the study protocol conformed to the ethical guidelines of the 1975 Declaration of Helsinki as reflected in a priori approval by CUIMC Institutional Review Board.

### Sequence analysis and variant annotation

Sample preparation, target-enrichment, sequencing process, read alignment, and variant calling were previously published^30,31^. We focused on variants that were predicted to have at least moderate to strong biological effects toward protein function and excluded those in intergenic and promoter regions. We used stringent quality filters and removed potential technical false-positive insertions and deletions (indels) using ATAV as previously described ^30,32^. We excluded variants failing quality cutoffs in gnomAD or those identified as sequencing artifacts through a comparison of in-house control sequencing data. Current guidelines recommend considering all variants with a minor allele frequency (MAF) of less than 1% at the population level. Thus, we filtered the variants based on the overall MAF of less than 1% in the Genome Aggregation Database (gnomAD)^33^. Variants previously reported as pathogenic were identified using the HGMD and ClinVar. We included only those annotated as pathogenic/likely pathogenic (P/LP) in ClinVar or disease-causing mutation (DM) in HGMD without any conflicting evidence within each database. In addition, we identified novel protein-truncating variants (PTVs) not previously reported in either HGMD or ClinVar. As the initial yield of individuals carrying candidate pathogenic variants was significantly higher than expected, we employed a stringent filter by inheritance mode and subpopulation MAF based on the data from gnomAD and Exome Aggregation Consortium (ExAC): MAF ≤ 10^−4^ for dominant disorders and MAF ≤ 10^−3^ for recessive disorders^30,34,35^. We used Loss-Of-Function Transcript Effect Estimator (LOFTEE) filter to exclude PTVs with a false prediction. A detailed description of genetic terminology in this study has been described previously^30^.

### Manual variant classification and clinical data review

Two independent genetic analysts performed a first-tier, stringent analysis of the CLD cohort to reach a consensus classification according to the ACMG-AMP guidelines^36^. We next performed a second-level manual curation of the CLD cohort using lower stringency filters, which identified several well-defined pathogenic variants that were excluded because they either have a MAF above 1% in some ethnic subpopulations or did not pass the stringent sequencing quality filters. This procedure had been successfully used to increase diagnostic yield in prior studies^13,37^. Subsequently, a multidisciplinary group of experts, including genetic counselors, geneticists, molecular pathologists, and clinicians, reviewed the available clinical information in individuals carrying P/LP variants to detect phenotypic concordance with the associated mode of inheritance of disease. If diagnostic evidence was insufficient based on chart review, a follow-up plan was recommended to clarify the significance of the genetic findings.

### Statistical analysis

We compared the probability of being loss-of-function intolerant (pLI) and Z scores for genes using an analysis of variance (ANOVA) test to compare differences between the three groups. We analyzed the clinical variables between those with and those without a genetic diagnosis using the Chi-squared test. All statistics and genetic analyses were done in R statistical software (Version 4.0.0). A p-value of <0.05 was considered significant after correction for multiple hypothesis testing.

## Supporting information

Gene list

Variant list

## Data Availability

The data sets supporting this study are available at the following publicly accessible websites: igmdx.org and atavdb.org. All data produced in the present study are available upon reasonable request to the authors.

https://atavdb.org

## List of Abbreviations

ES: whole-exome sequencing
CLD: chronic liver disease
CKD: chronic kidney disease
HGMD: Human Gene Mutation Database
PTV: protein truncating variants
MAF: minor allele frequency
GWAS: next-generation sequencing: NGS, genome-wide association studies
ACMG-AMP: American College of Medical Genetics and Genomics and the Association for Molecular Pathology
OMIM: Online Mendelian Inheritance in Man database
HPO: Human Phenotype Ontology
gnomAD: Genome Aggregation Database
ExAC: Exome Aggregation Consortium
AD: autosomal dominant
AR: autosomal recessive
XLD: X-linked dominant
XLR: X-linked recessive disorders
DM: disease-causing mutation
P/LP: pathogenic/likely pathogenic
CADD: Combined Annotation Dependent Depletion
QC: quality control

## Figure and table legends

Table 1: A comparison of clinical characteristics for those with or without a genetic diagnosis in the liver disease cohort.

Table 2: Hemochromatosis, alpha1 antitrypsin deficiency, and genetic diagnosis in the liver disease cohort. High MAF candidate pathogenic variants in *HFE* and *SERPINA1*. # Individuals with a homozygous pathogenic variant or two heterozygous pathogenic variants reported in HGMD or ClinVar. ^*^Unable to determine the phase of two variants. ^$^: cases with sufficient clinical evidence of liver phenotypes consistent with the genetic diagnosis. One case with *H63D* has liver phenotypes consistent with hemochromatosis.

## Acknowledgments

thank all the participants for enrolling into first studies. The study was supported by the Columbia precision medicine initiative.

