## Supplementary material for "Challenges associated with diagnostic exome sequencing in liver diseases": S.fig 1

Supplementary Figure 1: The search and establishment of a Mendelian liver disease gene list

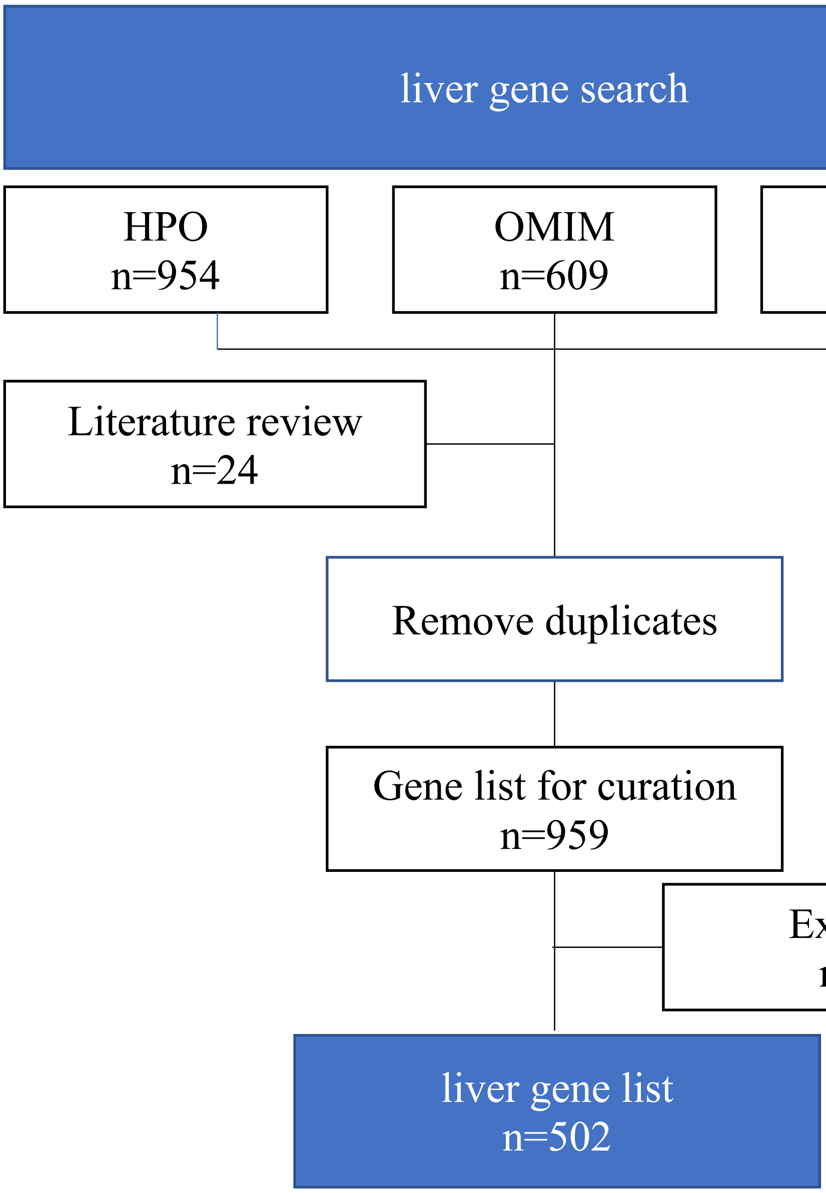


Keywords or phrase used to search for liver gene list: "liver", "biliary", "hepa", "cirrhosis", "Reye syndrome", "transaminases", "gallbladder", "cholestasis", "jaundice", "bile", "cholecystitis", "cholelithiasis", "portal", "steatosis", "dyslipidemia", "hyperlipidemia", "hypercholesterolemia", "hypertriglyceridemia", "insulin resistance", "hepatic fibrosis", "hypobetalipoproteinemia", "abetalipoproteinemia", "glycogen storage diseases", "lysosomal storage disorders", "lysosomal acid lipase deficiency", "fatty acid oxidation", "Gaucher’s", "lipodystrophies", "polycystic ovarian syndrome", "hemochromatosis",
