## Supplementary material for "Challenges associated with diagnostic exome sequencing in liver diseases": S.figure2

Supplementary figure 2. A stringent filtering and ACMG-AMP classification revealed an enrichement of pathogenic/likely pathogenic variants in CLD cohort

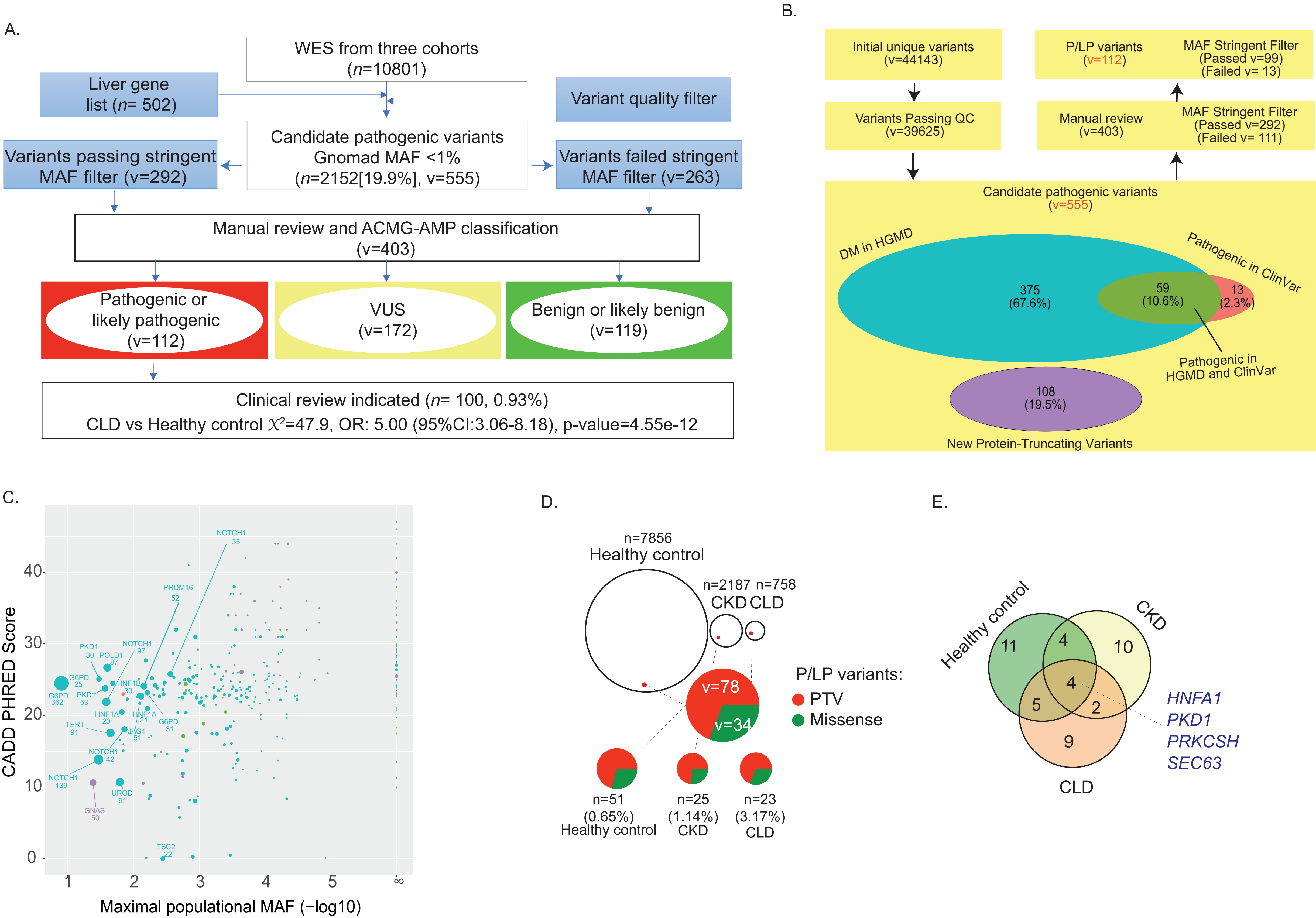
