## Supplementary material for "Challenges associated with diagnostic exome sequencing in liver diseases": S.table 1

Supplementary Table 1: The original research of the samples used in this study

| Original research | Current study population | n=10, 801 | |
|---|---|---|---|
| WHICAP | Population-based study of Medicare recipients aged ≥ 65 years in northern Manhattan | 2983 | 27.6% |
| Genetics of neurological disorders, OCD, ALS, Epilepsy, memory, schizophrenia | Self-reported healthy family members of patients with epilepsy, ataxia, neuromuscular disorders, brain malformations, neurodevelopmental disorders, OCD, schizophrenia, ALS, early-onset Parkinson disease, and other neurological disorder | 2893 | 26.8% |
| Diagnostic sequencing study of undiagnosed genetic disorders | Self-reported healthy family members of patients with undiagnosed disorders from general genetic clinic | 1564 | 14.5% |
| 27 additional studies | Self-reported healthy family members or control participants to a variety of genetic studies | 325 | 3.0% |
| **Genetic study of chronic liver diseases** | Self-reported healthy family members to chronic liver disease genetic study | 94 | 0.9% |
| Genetics study of kidney and Genitourinary disorders | Patients with chronic kidney diseases or genitourinary disorders | 2187 | 20.2% |
| **Genetic study of chronic liver diseases** | Patients with chronic liver diseases | 758 | 7% |

WHICAP: Washington Heights-Inwood Columbia Aging project; ALS: amyotrophic lateral sclerosis; OCD: obsessive compulsive disorder.
