## Supplementary material for "Challenges associated with diagnostic exome sequencing in liver diseases": S.table 2

Supplementary Table 2: Clinical characteristics and genetic testing information for three cohorts

| Characteristics | Healthy control cohort (n=7856) | Proportion in healthy control cohort | Chronic kidney disease cohort (n=2187) | Proportion in CKD cohort | Chronic liver disease cohort (n=758) | Proportion in CLD cohort |
|---|---|---|---|---|---|---|
| Gender | | | | | | |
| Male | 3176 | 40.4% | 945 | 43.2% | 351 | 46.3% |
| Female | 4680 | 59.5% | 1242 | 56.8% | 407 | 53.7% |
| Age at time of study entry | | | | | | |
| 0-21 yr | 240 | 3.1% | 278 | 12.7% | 255 | 33.6% |
| 22-44 yr | 1416 | 18.0% | 713 | 32.6% | 150 | 19.8% |
| 45-64 yr | 597 | 7.6% | 800 | 36.6% | 226 | 29.8% |
| ≥ 65 yr | 103 | 1.3% | 396 | 18.1% | 127 | 16.8% |
| Unspecified | 5500 | 70.0% | | | | |
| Self-declared race/ethnicity | | | | | | |
| White | 3427 | 43.6% | 1113 | 50.9% | 367 | 48.4% |
| Hispanic | 2303 | 29.3% | 435 | 19.9% | 136 | 17.9% |
| Black | 1070 | 13.6% | 330 | 15.1% | 95 | 12.5% |
| Asia | 420 | 5.3% | 224 | 10.2% | 61 | 8% |
| Other or unspecified | 636 | 8.1% | 85 | 3.9% | 99 | 13.1% |
| Sequencing Modality | | | | | | |
| Whole Exome Sequencing | 7735 | 98.4% | 2187 | 100% | 758 | 100% |
| Whole Genome Sequencing | 121 | 1.6% | | | | |
| Exome Capture Kit | | | | | | |
| Roche | 5954 | 76.7% | 1495 | 68.4% | 221 | 29.2% |
| IDTERPv1 | 1147 | 14.8% | 692 | 31.5% | 537 | 70.8% |
| Other | 664 | 8.6% | | | | |
