## Supplementary material for "Challenges associated with diagnostic exome sequencing in liver diseases": S.table 3

Supplementary Table 3: The inheritance mode and biological annotations of 502 genes related to Mendelian disorders and liver phenotypes

| 502 liver genes | Hepatocyte Intrinsic | Biliary | Metabolism | Immune | Development |
|---|---|---|---|---|---|
| AD (n=61) | 26 | 9 | 12 | 9 | 31 |
| AD& AR (n=62) | 26 | 12 | 26 | 16 | 33 |
| AR (n=363) | 161 | 53 | 180 | 60 | 224 |
| XLD (n=1) | 0 | 0 | 1 | 0 | 0 |
| XLR(n=15) | 6 | 2 | 3 | 5 | 10 |

AD: Autosomal dominant, AR: Autosomal recessive, XLD: X-linked dominant, XLR: X-linked recessive.
