## Supplementary material for "Challenges associated with diagnostic exome sequencing in liver diseases": S.table 4

Supplementary Table 4: Stepwise filtering to identify individuals with likely pathogenic variants in liver genes

| **Total WES (n=10,804)** | **Healthy control cohort (n=7856)** | **Kidney disease cohort (n=2187)** | **Liver disease cohort (n=758)** |
|---|---|---|---|
| Initial total variants count | 215,147 | 57,724 | 19,916 |
| Variants: QC filter | 186,818 | 50,443 | 17,884 |
| Unique variants | 31,669 | 14, 319 | 7,202 |
| Pathogenic HGMD or ClinVar | 1873 | 519 | 208 |
| New Protein Truncating Variants | 145 | 38 | 16 |
| Individuals carry candidate pathogenic variants | 1577 (20.1%) | 416 (19.0%) | 159(21.0%) |
